# Genetic variation supports a causal role for valproate in prevention of ischemic stroke

**DOI:** 10.1101/2023.02.14.23285856

**Authors:** Ernst Mayerhofer, Livia Parodi, Kaavya Narasimhalu, Stefan Wolking, Andreas Harloff, Marios K Georgakis, Jonathan Rosand, Christopher D Anderson

**Affiliations:** Center for Genomic Medicine, Massachusetts General Hospital, Boston, MA; Program in Medical and Population Genetics, Broad Institute of Harvard and the Massachusetts Institute of Technology, Boston, MA, USA; McCance Center for Brain Health, Massachusetts General Hospital, Boston, MA, USA; Department of Neurology, Brigham and Women’s Hospital, Boston, MA, USA; Department of Neurology and Epileptology, University Hospital Aachen, Germany; Department of Neurology and Neurophysiology, Medical Center – University of Freiburg, Faculty of Medicine, University of Freiburg, Germany; Institute for Stroke and Dementia Research (ISD), University Hospital, Ludwig-Maximilians-University (LMU) Munich, Munich, Germany

**Author notes:** Corresponding author Christopher D Anderson, Brigham and Women’s Hospital, Boston.

## Abstract

Valproate is a candidate for ischemic stroke prevention due to its anti-atherosclerotic effects in vivo. Although valproate use is associated with decreased ischemic stroke risk in observational studies, confounding by indication precludes causal conclusions. To overcome this limitation, we applied Mendelian randomization to determine whether genetic variants that influence seizure response among valproate users associate with ischemic stroke. We derived a genetic score for valproate response using genome-wide association data of seizure response after valproate intake from the Epilepsy Pharmacogenomics Consortium. We then tested this score among valproate users of the UK Biobank for association with incident and recurrent ischemic stroke using Cox proportional hazard models. Among 2,150 valproate users (mean 56 years, 54% females), 82 ischemic strokes occurred over a mean 12-year follow-up. Higher valproate response genetic score was associated with higher serum valproate levels (+5.78 µg/ml per one SD, 95% CI [3.45, 8.11]). After adjusting for age and sex, higher valproate response genetic score was associated with lower ischemic stroke risk (HR per one SD 0.73, [0.58, 0.91]) with a halving of absolute risk in the highest compared to the lowest score tertile (4.8% vs 2.5%, p-trend=0.027). Among 194 valproate users with prevalent stroke at baseline, a higher valproate response genetic score was associated with lower recurrent ischemic stroke risk (HR per one SD 0.53, [0.32, 0.86]) with reduced absolute risk in the highest compared to the lowest score tertile (3/51, 5.9% vs. 13/71, 18.3%, p-trend=0.026). The valproate response genetic score was not associated with ischemic stroke among the 427,997 valproate non-users (p=0.61), suggesting minimal pleiotropy. In an independent cohort of 1,241 valproate users of the Mass General Brigham Biobank with 99 ischemic stroke events over 6.5 years follow-up, we replicated our observed associations between the valproate response genetic score and ischemic stroke (HR per one SD 0.77, 95% CI: [0.61, 0.97]). These results demonstrate that a genetically predicted favorable seizure response to valproate is associated with higher serum valproate levels and reduced ischemic stroke risk among valproate users, providing causal support for valproate effectiveness in ischemic stroke prevention. The strongest effect was found for recurrent ischemic stroke, suggesting potential dual-use benefits of valproate for post-stroke epilepsy. Clinical trials will be required in order to identify populations that may benefit most from valproate for stroke prevention.

## Introduction

Valproate is a widely used antiepileptic drug that has been associated with decreased risk for ischemic stroke in observational studies.^1–4^ Valproate is assumed to exert this preventive effect by inhibiting histone deacetylase 9 (HDAC9),^5, 6^ which in animal studies has been found to lead to a stabilizing and anti-inflammatory effect on atherosclerotic plaques.^7–9^ Although genetic variants in the *HDAC9* gene have been repeatedly and robustly associated with large-artery stroke in population-based and case-control genome-wide association studies (GWAS),^10, 11^ causal evidence supporting a role for valproate in stroke prevention through this mechanism in humans is still missing. One of the reasons for this gap is that observational studies are prone to biases and thus cannot deliver evidence for a causal drug effect.^12^ While randomized clinical trials provide the needed evidence, they are often tailored to a specific indication and can be underpowered for secondary endpoints or uncommon side effects,^13^ making efficient evaluation of drug repurposing challenging.

Germline genetic variation, constant throughout the lifespan and thus not prone to confounding, can be leveraged to assess whether a drug causally contributes to a specific outcome. If there are genetic variants that are known to influence drug response, the drug users in an observational study can be divided according to the predicted genetic response. Because the prescribing health care providers and the patients are not aware of these genetic variants at the time of prescription, the cohort of drug users can be considered randomized and blinded by genetics. If the genetic variants that predispose to better drug response are associated with the outcome of interest in users of the drug, the results support the hypothesis that the drug causally contributes to the outcome. Pleiotropic effects of the genetic variants that have an independent effect on the outcome can be ruled out if no associations are found among non-drug users. We have recently applied this form of *in silico* simulation of a randomized controlled trial, a special case of Mendelian randomization, to show that statins causally contribute to intracerebral hemorrhage.^14^

Utilizing this framework, we investigated the causal contribution of valproate use on risk of incident and recurrent ischemic stroke. To gain insight into clinically relevant effects on atherosclerosis, we additionally investigated the effect on myocardial infarction. We leveraged data from a previously published GWAS of clinical response to three antiepileptic drugs (valproate, lamotrigine, and levetiracetam)^15^ to construct and validate a genetic score for valproate response among valproate users in the UK Biobank (UKB) and study its association with the selected outcomes (**Figure 1A**). To rule out the possibility that detected associations are driven by an antiepileptic drug class effect, we also used the same approach to study the effects of lamotrigine and levetiracetam on ischemic stroke. To assess the robustness of our findings, we replicated observed associations in an independent cohort of valproate users of the Mass General Brigham Biobank (MGBB).

**Figure 1.**
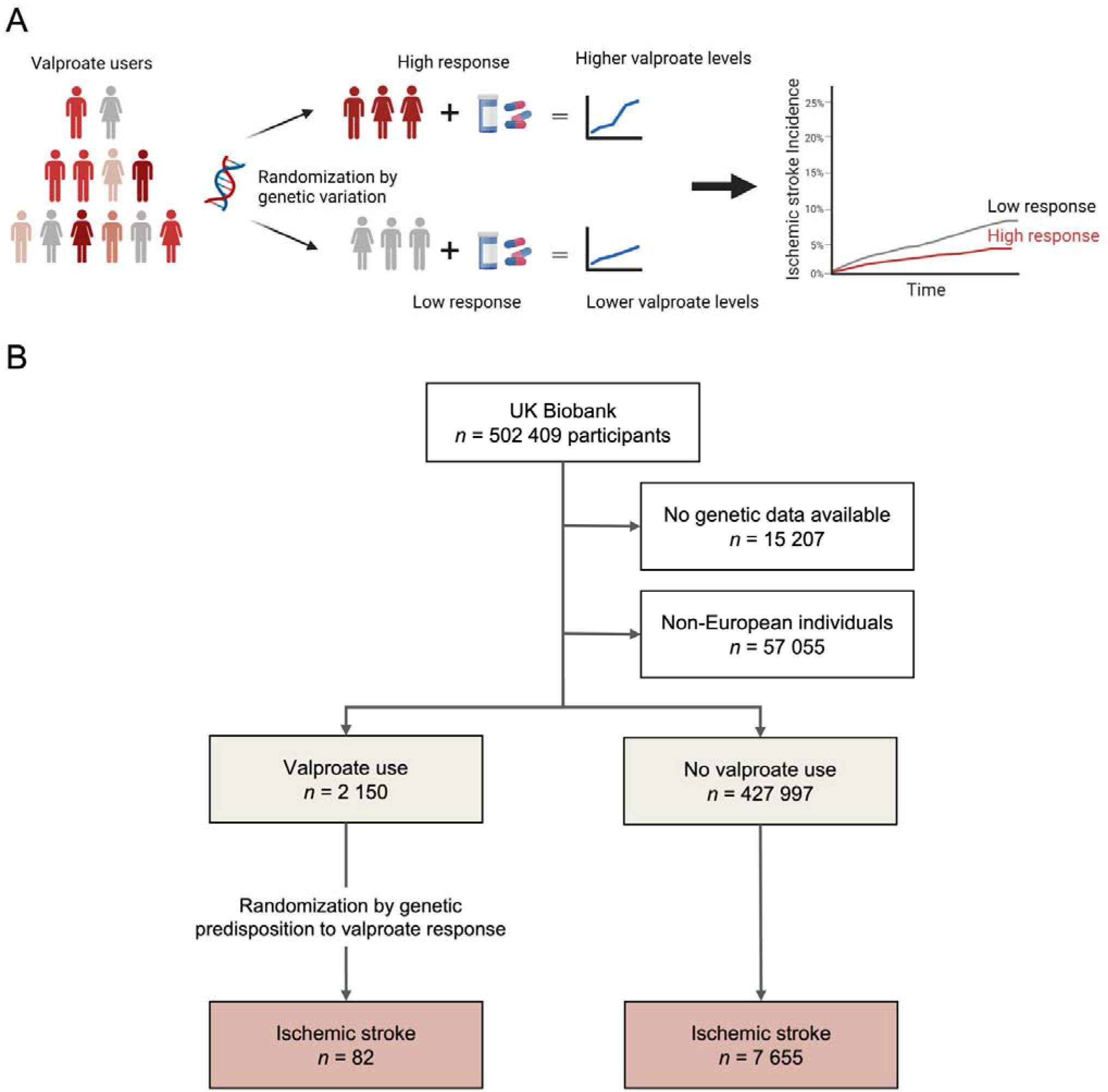
Study overview. **A:** Concept of the *in silico* trial randomized by genetic variation. A genetic score consisting of genetic variants known to predispose to higher likelihood of seizure freedom after valproate intake was associated with serum valproate levels and incidence of ischemic stroke. **B:** Study flow. After exclusion of Non-European individuals and those without genetic data, 2,150 valproate users and 427,997 valproate non-users were identified.

## Methods

### Study population

The UKB is an ongoing, population-based prospective cohort study of over 500,000 individuals aged 37-73 years at baseline, recruited from 2006-2010 in 22 assessment centers across the UK.^16^ A wide range of baseline data including phenotyping assessments, biochemical assays, genome-wide genotyping, and primary care data (in a subset of the total cohort) together with ongoing follow-up outcome data is available. Only individuals with available genetic data were included in the present study. Because we used genetic variants^15^ and a linkage disequilibrium reference panel^17^ that were both discovered only in individuals of European ancestry (see below), we further restricted our cohort to European ancestry participants.

The UKB has institutional review board approval from the Northwest Multi-Center Research Ethics Committee (Manchester, UK). All participants provided written informed consent. We accessed the data following approval of an application by the UKB Ethics and Governance Council (Application No. 36993).

### Mendelian randomization approach

Because the genetic variants that are used for the exposure (in our study, valproate, lamotrigine, and levetiracetam clinical response) are derived from a GWAS including only individuals exposed to those medications, but stroke and myocardial infarction outcome GWAS have been performed among drug users and non-users, we used an individual-level approach in the UK Biobank to test for drug-specific effects and assess for pleiotropic effects of our genetic instruments. We constructed a genetic score for response to each drug and tested each score for association with the outcomes of interest among individuals exposed or not exposed to each medication. Because of the random assortment of common alleles in a population, genetically predicted drug response is randomly allocated, and thus an association of the genetic response score with the outcome of interest in those exposed to the drug provides evidence of a causal drug effect. Further, pleiotropic effects of the genetic variants that inadvertently modify the risk for chosen outcomes independent of the drug can be ruled out if there is no association among individuals not exposed to the drug. This approach has been described by us and others in previously published work.^14, 18^

### Identification of antiepileptic drug users

We identified valproate, lamotrigine, and levetiracetam drug users via verbal baseline interview and primary care prescription data, allowing us to capture drug prescriptions within a time period between 1978 to 2018. We have previously reported the details of our pipeline to extract medication data from UKB primary care data.^14^ First, all available ever-approved formulations for valproate, lamotrigine, and levetiracetam were gathered by using international nonproprietary (INN) names, former and current trade names in the UK (via the National Health Service Dictionary of Medicines and Devices [DM+D] browser, https://services.nhsbsa.nhs.uk/dmd-browser/search), and their associated DM+D and British National Formulary (BNF) codes (**Supplemental Table S1**). Then, all primary care prescription data and the verbal interview data were searched for these formulations. Individuals were considered as users of a drug if they reported intake of one of the formulation names containing the drug at any verbal interview (baseline or follow-up, UKB field 20003) or if they had two or more prescriptions of a formulation containing the drug in the primary care data (gp_scripts table). The first drug prescription date for each individual was defined either as the first prescription date from primary care data or as the verbal interview date, whichever was earlier and available. To assess whether patients were on monotherapy or had other antiepileptic drugs prescribed, we extracted prescriptions for the most common antiepileptic drugs (**Supplemental Table S1**) between first prescription of valproate and end of follow-up.

### Construction of the genetic scores for antiepileptic drug response

We used genome-wide association data from the Epilepsy Pharmacogenomics Consortium (EpiPGX) on seizure freedom after antiepileptic drug intake in European ancestry patients with generalized epilepsy.^15^ In that study, participants were defined as treatment responder if they were seizure free under continuous treatment for at least one year, and as treatment non-responders if they had 50% of pretreatment seizure frequency or higher under adequate dosing of the drug according to a specialist.^15^ The cohort included patients on valproate (n=565), lamotrigine (n=387), and levetiracetam (n=209).^15^ Association tests were performed based on responder vs. non-responder status for each of the drugs.^15^ There was no participant overlap between the EpiPGX GWAS and the UKB. Association results were available for single nucleotide polymorphisms (SNPs) associated with drug response at p<0.05 (n=162,242 for valproate, n=162,666 for lamotrigine, and n=162,430 for levetiracetam).

To construct the genetic scores to be used as instruments for valproate, lamotrigine, and levetiracetam response, we leveraged PRS-CS (polygenic prediction via bayesian regression and continuous shrinkage priors), a novel unsupervised polygenic prediction method for that uses a high-dimensional Bayesian regression framework to derive a genetic score from GWAS summary statistics without requiring an external validation cohort.^17^ PRS-CS takes linkage disequilibrium of genetic variants into account by using an external linkage disequilibrium reference panel and outperforms traditional clumping and thresholding approaches such as PRSice.^17, 19^ We used PRS-CS with default parameters to generate SNP weights for response to each drug, which yielded weights for 33,089, 33,300, and 32,736 SNPs for valproate, lamotrigine, and levetiracetam, respectively.

To test robustness of the discovered associations, we performed sensitivity analyses with an alternative genetic instrument for valproate response that was derived using a clumping and thresholding approach. Following previously described approaches for drug response Mendelian randomization studies,^14, 20^ we selected the SNPs from the EpiPGX GWAS results that were associated with valproate response at *p*<5x10^-5^ and clumped at r^2^<0.001 based on the 1000 Genomes European reference panel.^20^ The alternative genetic score for valproate response consisted of 20 SNPs that were retained after clumping of 139 SNPs.

Finally, using the weights derived from PRS-CS and clumping and thresholding, we calculated the individual genetic scores for corresponding drug users in the UKB using imputed genotype data. For assessment of appropriate randomization, Kaplan-Meier curves, and calculation of absolute risk differences, individuals were divided in genetic score tertiles. The SNPs and weights for the genetic scores are provided in **Supplemental Tables S2-S5**.

### Validation of the genetic score on serum valproate response to normalized valproate dosing

We aimed to test the effect of the derived genetic score for valproate response on valproate serum levels to confirm its validity to test the hypothesis whether valproate has an effect on ischemic stroke through serum level-dependent effects. Genetic variants that are associated with seizure response to valproate could be unrelated to the effect of valproate on ischemic stroke, if they influence a pathway that is not related to drug metabolism, but rather further downstream in its effect on seizure prevention. However, if the genetic variants predict valproate response through an effect on valproate serum levels below the threshold of impacting prescriber behavior, they are a proxy for genetically predicted drug exposure and thus can be used as an instrument for randomization in the test for an effect on ischemic stroke. In this special case of Mendelian randomization, this is assertion of the relevance assumption of the genetic variants.

Valproate serum levels were gathered from the primary care clinical data by using the Read codes ‘44W4.’ and ‘XE25d’. Values that were 0 (indicating off-valproate situations) and those higher than 200 µg/ml (potentially erroneous or not in µg/ml) were discarded. For each serum level value, the taken valproate dose at the time of measurement was approximated. First, the duration in days between the prescriptions before and after the date of the serum level measurement was calculated. Then, the quantity of tablets was multiplied by the dose of each tablet, divided by the duration of the prescription interval, yielding the average daily dose in mg per day. The association of valproate dose with valproate serum levels was tested in a linear regression model with valproate serum level as dependent variable and average daily valproate dose and the genetic score as independent variables. The model was additionally adjusted for age at the time of serum level measurement and sex. Levels for lamotrigine or levetiracetam were not available in the UK Biobank.

### Outcome ascertainment

UKB participants’ records have been linked with inpatient hospital codes, primary care data, and death registry for longitudinal follow-up. Outcome events were gathered from hospital admissions and death registry data using International Classification of Diseases (ICD) 10 codes that were aligned with the diagnostic algorithm in the UKB (https://biobank.ndph.ox.ac.uk/showcase/ukb/docs/alg_outcome_main.pdf). Incident events were defined as events occurring after baseline and after the first drug prescription date. Because only very limited information on ischemic vs. hemorrhagic stroke subtypes before baseline exists in the UKB, recurrent ischemic strokes were defined as ischemic strokes occurring in individuals with a history of any stroke at baseline, as defined in the UKB field 42006. The ICD-10 codes used for ascertainment of the outcomes are supplied in **Supplemental Table S6.** Stroke outcomes in the UKB have been routinely used in genetic association studies, including the most recent GWAS of stroke risk.^11^

### Association of the genetic scores with outcomes

To explore the effects of valproate on the selected outcomes, cause-specific Cox proportional hazard models censored for death were used with the valproate-specific genetic scores as independent variable, adjusted for age, sex, principal components (PC) 1-3, and genotyping assay in the cohort of valproate users. Although ischemic stroke subtypes are not available in the UKB, we tried to investigate the pathophysiological mechanism of valproate’s action on stroke prevention. To approximate cardioembolic stroke, ischemic stroke in the setting of atrial fibrillation was analyzed by adding an interaction term of the genetic score with prevalent atrial fibrillation in the model for ischemic stroke and performing subgroup analyses in individuals with and without a diagnosis of atrial fibrillation before or within six months after ischemic stroke. Because of the small cohort size, the main analyses were performed in all valproate users regardless of potential cryptic relatedness, and sensitivity analyses were performed in a cohort restricted to unrelated individuals (KING kinship coefficient < 0.0884). Chi-Square test for trend in proportions were used to assess significant differences in absolute stroke risk between genetic score tertiles. To rule out that observed associations between the genetic scores and the outcomes are caused by pleiotropic effects of the genetic score not related to valproate use, we tested the same associations among non-valproate users, thus assessing the independence and exclusion restriction assumptions of Mendelian randomization. To further rule out an antiepileptic drug class effect, all analyses were repeated with the genetic scores for lamotrigine and levetiracetam response among users of the respective drugs. To exclude drug interactions, we also performed a sensitivity analysis among the cohorts of patients on valproate monotherapy.

### Replication analyses

We aimed to confirm our discoveries in the MGBB, an ongoing prospective clinical research cohort of patients of Mass General Brigham (MGB), the parent organization of Massachusetts General Hospital (MGH) and Brigham and Women’s Hospital (BWH) in Boston, Massachusetts, USA. All patients aged 18 years or older presenting to any of the MGB clinics consenting to broad research are included.

Patients are recruited in-person at MGH and BWH and online through an electronic patient gateway. The MGBB provides ongoing electronic health record data with ICD codes for outcomes, imaging, and, on a subset of individuals, genetic data. Recruitment has been ongoing since 1998 and to date, more than 133,000 patients have been included in the Biobank and over 65,000 have been genotyped. Genotyping has been performed in batches on different arrays, with batches 1-9 performed by MGH and batches 10-13 by the Broad Institute. To minimize batch effects and variation across different genotyping arrays, many of the individuals that have been genotyped in batches 1-9 have been re-genotyped in batch 13. For the current analyses, we considered only individuals that were genotyped in batches 10-13, which we combined and imputated together on the Michigan Imputation Server using the Haplotype Reference Consortium v.1.1 reference panel. We used exactly the same PRS-CS approach to construct the genetic score in MGBB participants.

We queried the MGB Biobank database using the same criteria that we used in the UKB. We identified patients with available genetic data and two or more oral prescriptions of valproate. Most of the prescriptions contained instruction texts from which we could extract the daily dose by multiplying the medication dose by the prescribed number of times taken daily. Outcomes were identified using inpatient ICD-9 and ICD-10 codes (**Supplemental Table S6**) and were classified as prevalent or incident events regarding their occurrence before or after the date of the first valproate prescription. We used the same statistical analysis approaches for the replication analyses in the MGBB as in the UKB. Linear regression models were used to assess the associations of the average valproate dose and the genetic score with valproate serum levels. For the association of the genetic score with outcomes, Cox proportional hazard models were constructed with the time to event as the number of days between the date of the first valproate prescription and incident event date for patients for which an outcome occured, and the number of days between the first valproate prescription and the last encounter for patients for which no outcome occured. We detected significant associations between the principal components and age and sex, most likely due to chance imbalance across genotyping batches; people in later batches were better powered and consisted of older individuals and more women, and thus models adjusted for age and sex and the principal components introduced collinearity to our model. Because we found no associations between the genetic score for valproate response with age and sex, indicating near-perfect randomization, we removed age and sex from our Cox models and the reported effect estimates are from models with the genetic score as main predictor, adjusted for principal components 1-3 which still contain age and sex information.

### Software and statistical methods used

ANOVA and Chi-Square test were used for comparison of continuous and discrete baseline variables, respectively. SNP extraction and genetic score calculation was performed with PLINK and bcftools, relationship inference was performed with KING.^21–24^ Data extraction, curation, preparation, statistical analysis, and figure generation was done with RStudio on Mac OS X.^25^ This study followed the guideline for the usage of Mendelian Randomization in observational studies (STROBE-MR).^26^

### Data availability

UK Biobank participant data are available through the UK Biobank after approval of a research proposal. The SNPs used for construction of the genetic scores were obtained from the authors of the original publication.^15^

## Results

### Baseline characteristics

We identified 2,150 valproate users after exclusion of non-European ancestry individuals and those with unavailable genetic data (**Figure 1B**, **Table 1**). The date range for the prescriptions was July 1987 to March 2018, and the mean time between first and last prescription was 6.5 ± 6.9 years. Valproate users had a significant higher rate of vascular risk factors compared to valproate non-users, but no difference in the genetic score (**Table 1**). The genetic score for valproate response was normally distributed among valproate users (**Figure 2A**). When comparing valproate users stratified by genetic score, no differences in baseline characteristics, cardiovascular risk factors, antiplatelet and statin use, or approximated valproate doses (among the 1,387 individuals with available prescription data) were found across genetic score tertiles, indicating an appropriate randomization (**Table 2**). Most patients were on valproate monotherapy, and all tertiles had average valproate serum levels in the therapeutic range (**Table 2**).

**Figure 2.**
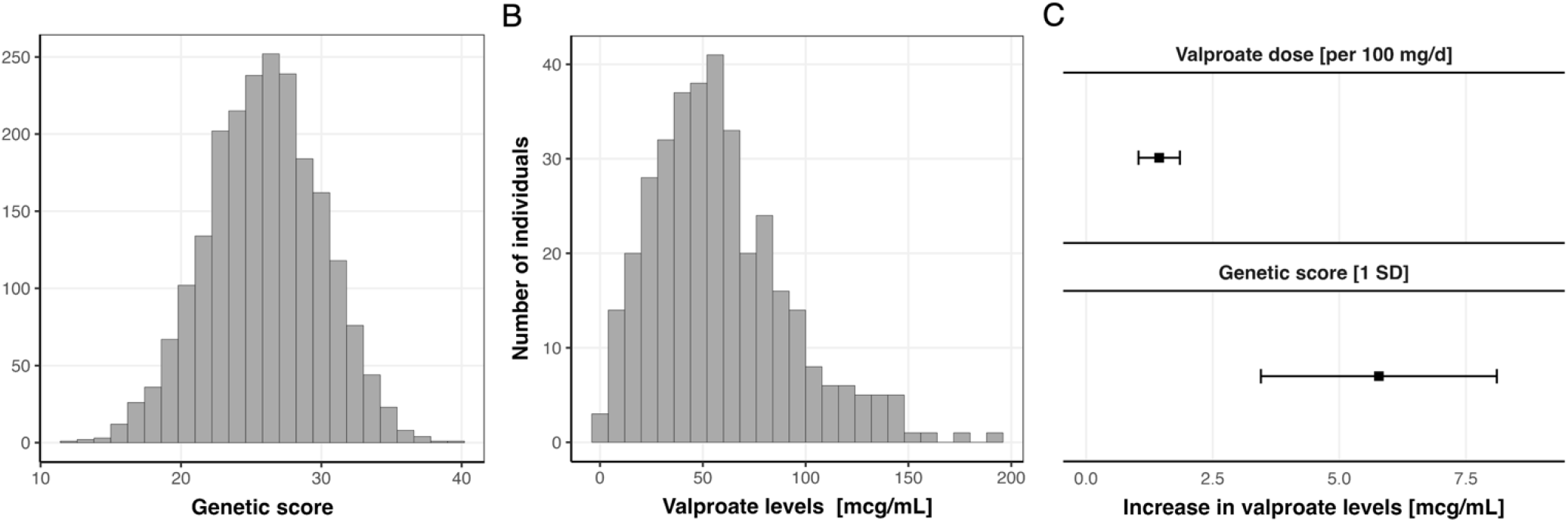
Valproate serum levels and their association with the genetic score for valproate response in the UK Biobank. **A**: Distribution of the genetic score among the 2,150 valproate users. **B:** Distribution of the 549 valproate serum level values among 202 valproate users. **C:** Association of approximated daily valproate dose and the genetic score for valproate response with valproate serum levels in a linear regression model adjusted for age and sex. Depicted in the top is the coefficient for valproate dose, on the bottom it is the genetic score for valproate response, suggesting that the genetically predicted valproate response acts through valproate metabolism.

**Table 1.** Baseline characteristics of valproate users compared to valproate non-users.

|  | Valproate users<br>(N=2,150) | Valproate non-users<br>(N=427,997) | p value |
| --- | --- | --- | --- |
| Age at baseline, years, mean (SD) | 56.4 (8.18) | 56.9 (8.00) | 0.0106 |
| Female sex, n (%) | 1,153 (53.6) | 231,608 (54.1) | 0.667 |
| Hypertension, n (%) | 641 (29.8) | 113,259 (26.5) | <0.001 |
| Diabetes, n (%) | 156 (7.3) | 20,138 (4.7) | <0.001 |
| Hypercholesterolemia, n (%) | 335 (15.6) | 52,795 (12.3) | <0.001 |
| Smoking status |  |  | <0.001 |
| Never Smoker, n (%) | 1,111 (51.7) | 232,202 (54.3) |  |
| Ex-Smoker, n (%) | 702 (32.7) | 150,730 (35.2) |  |
| Current Smoker, n (%) | 328 (15.3) | 43,551 (10.2) |  |
| History of stroke, n (%) | 198 (9.2) | 6423 (1.5) | <0.001 |
| History of myocardial infarction, peripheral artery disease, or stroke, n (%) | 314 (14.6) | 27157 (6.3) | <0.001 |
| Statin use at baseline, n (%) | 509 (23.7) | 70,515 (16.5) | <0.001 |
| Antiplatelet use at baseline, n (%) | 428 (19.9) | 60,863 (14.2) | <0.001 |
| Genetic score for valproate response, mean (SD) | 25.9 (4.07) | 25.9 (4.17) | 0.85 |

**Table 2.**
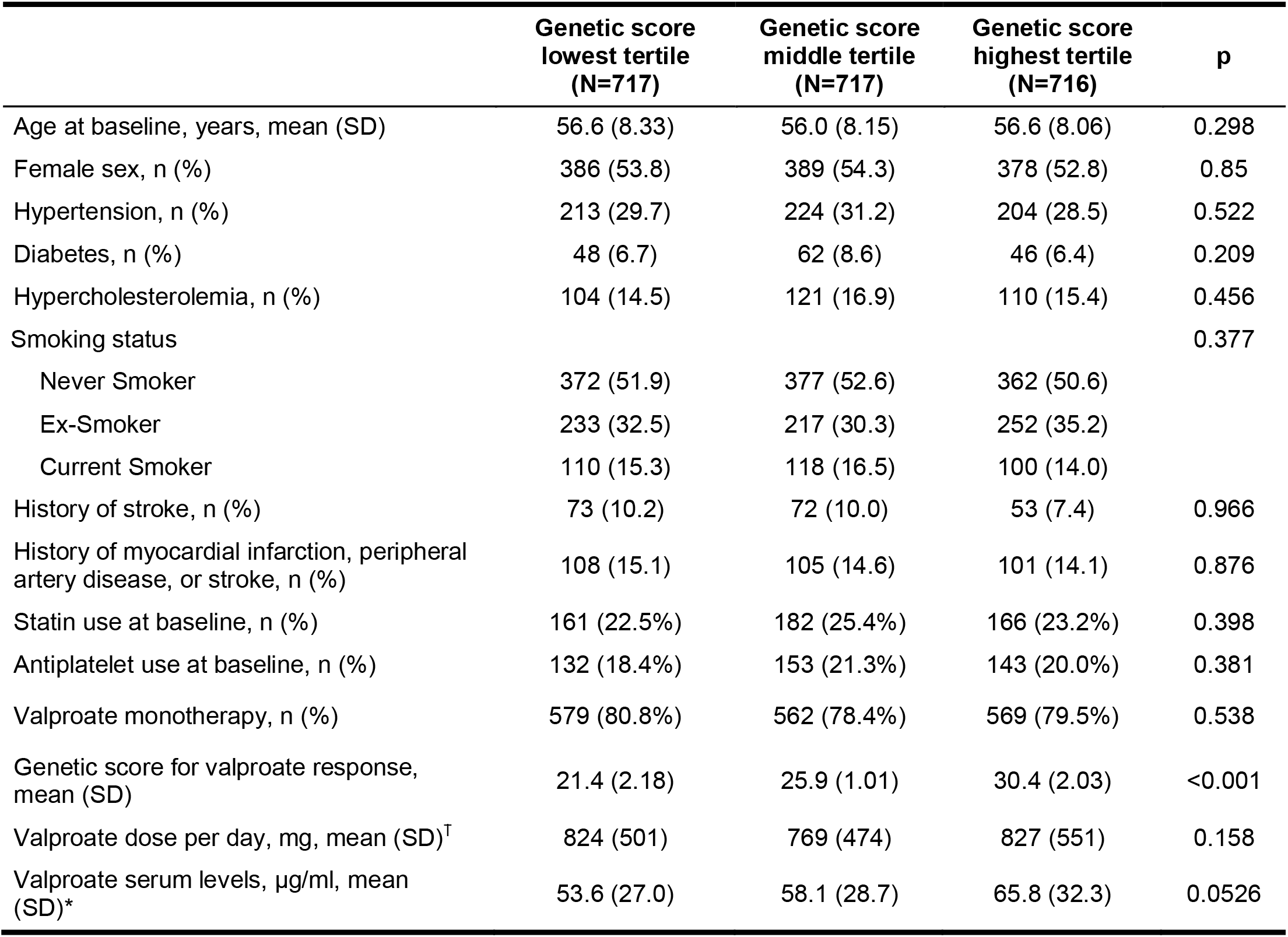
Baseline characteristics of valproate users stratified by the genetic score for valproate response. Higher scores indicate higher genetic predisposition for seizure freedom after valproate intake. ^†^ Approximated valproate doses were available for 1,387 individuals * 549 valproate serum levels were available for 202 individuals.

### Validation of the genetic score on serum valproate response to normalized valproate dosing

A total of 549 valproate serum level values were available for 202 valproate users after exclusion of 137 values that were zero and 41 values that were higher than 200 (**Figure 2B**). The approximated daily valproate dose was significantly associated with valproate serum levels (1.25 μg/ml per 100mg/day, 95% CI [0.85, 1.66], **Figure 2C**). We found a significant association of the genetic score with valproate serum levels, indicating higher average serum levels in participants with higher genetic scores (+5.78 µg/ml per one SD, 95%CI[3.45, 8.11], **Figure 2C**). These associations were replicated with the alternative genetic score (**Supplemental Table S7**). Serum levels for lamotrigine or levetiracetam were not available for validation of genetic scores.

### Association of the valproate genetic instrument with incident ischemic stroke

Among the 2,150 valproate users, 82 ischemic strokes occurred over a mean follow-up of 11.6 years. In Cox proportional hazard models, a higher genetic score for valproate response was associated with a lower risk of incident ischemic stroke (HR 0.73, 95% CI [0.58, 0.91] per one SD increase, **Figure 3A**). Individuals in the lowest tertile of the genetic score had an almost two-fold increased absolute risk for ischemic stroke compared to those in the highest score tertile (4.8% vs 2.5%, p-trend = 0.027, **Figure 3B**). Sensitivity analyses confirmed robustness of the findings among the 1,967 unrelated individuals (HR 0.75, 95% CI [0.59, 0.95] for 76 events, **Supplemental Table S8**) as well as with the alternative genetic score for valproate response using a clumping and thresholding approach (HR 0.75, 95% CI [0.59, 0.95], **Supplemental Table S9**). Restricting our cohort to the patients on valproate monotherapy (n=1,675), the associations were replicated but with wider confidence intervals due to lower power (**Supplemental Table S10**). We found no associations between the genetic scores for lamotrigine and levetiracetam response and ischemic stroke in the 1,108 lamotrigine and 789 levetiracetam users, respectively (p=0.76 and p=0.24, **Supplemental Tables S11-S12**).

**Figure 3.**
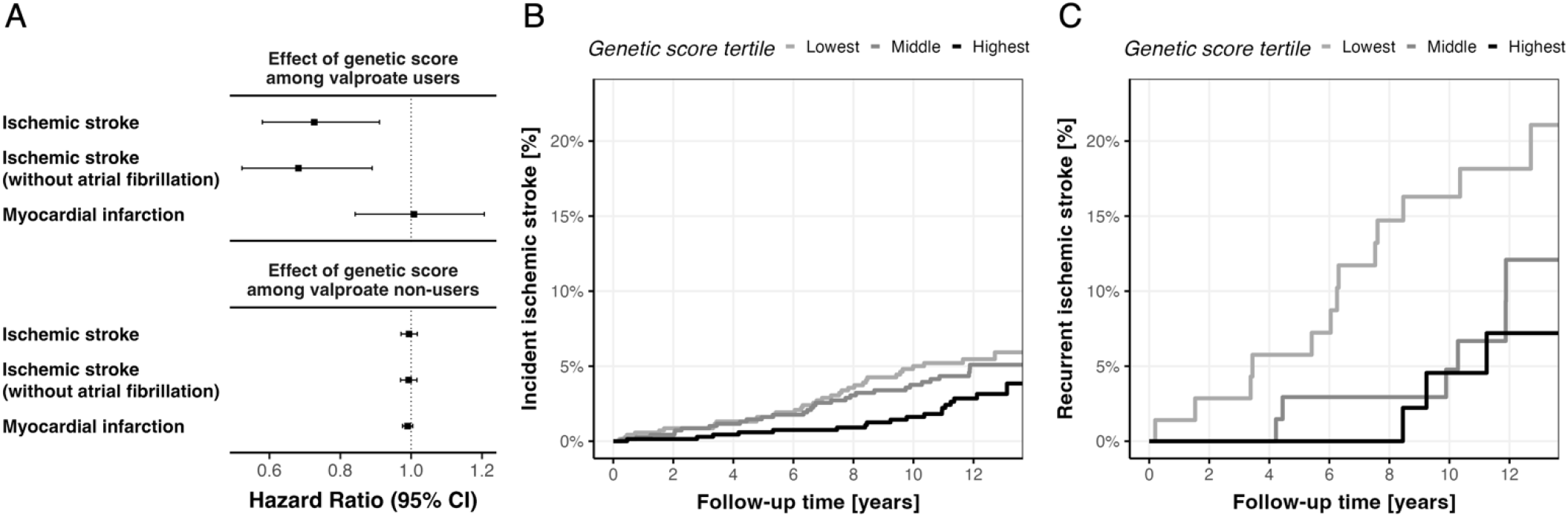
Associations between the genetic score for valproate response with outcomes in the UK Biobank. **A:** Hazard ratios of valproate on ischemic stroke, ischemic stroke without concurrent diagnosis of atrial fibrillation, and myocardial infarction. **B and C:** Kaplan-Meier plots for incidence of **(A)** 82 ischemic strokes among the 2,150 valproate users and **(B)** 22 recurrent ischemic stroke among the 194 valproate users with prevalent stroke at baseline, stratified by genetic score tertiles.

In the cohort of valproate users, 67 individuals had a diagnosis of atrial fibrillation before or within six months after ischemic stroke, and 24 ischemic strokes occurred among them. No interaction between prevalent atrial fibrillation and the genetic score was found (p=0.40). No association between the genetic score and ischemic stroke was found in this subgroup (p=0.51), however when removing the 67 individuals with atrial fibrillation from the cohort of valproate users, the association of the genetic score with incident ischemic stroke was more pronounced (HR 0.68, 95% CI [0.52, 0.89], **Figure 3A**), suggesting a better utility of valproate for prevention of ischemic stroke not related to atrial fibrillation.

### Association of the genetic score with recurrent ischemic stroke

Among the 194 individuals with prevalent stroke at baseline, 22 recurrent ischemic strokes occurred over a mean follow-up of 11.2 years. In Cox proportional hazard models, a higher genetic score was associated with decreased risk for recurrent ischemic stroke (HR 0.53, 95% CI [0.32, 0.86] per one SD increase). Although only few cases in total, individuals in the lowest tertile of the genetic score had a three-fold higher absolute risk for ischemic stroke compared to those in the highest tertile (13/71, 18.3% vs 3/51, 5.9%, p-trend=0.026, **Figure 3C**).

### Associations of the genetic score with myocardial infarction

No associations of the genetic score for valproate response were found for myocardial infarction (133 events, p=0.93) among valproate users. Also, no associations were found among valproate non-users, reassuring that our genetic instruments were not affected by horizontal pleiotropy (p=0.18, **Figure 3A**).

### Replication in the MGB Biobank

We identified 1,241 valproate users with available genetic data in the MGBB, among which 99 ischemic stroke events and 126 myocardial infarction events occurred over a median follow-up of 6.7 years. In this independent cohort we found 839 patients with 6,353 valproate serum level measurements. The associations between prescribed valproate dose and the genetic score with valproate serum levels replicated with almost identical effect estimates (**Supplemental Table S13**). We also found significant association between the genetic score for valproate response and ischemic stroke (HR 0.77, 95% CI: [0.61, 0.97]), and no association between the genetic score and myocardial infarction (p=0.68).

## Discussion

In this study, we used common genetic variants to stratify 2,150 valproate users in the UKB by their genetically predicted response to valproate and investigated their risk of incident ischemic stroke and myocardial infarction. We leveraged data from a GWAS of seizure control after valproate intake from a cohort of patients with epilepsy and applied it to medication prescription and intake data of 502,000 individuals from a population-based observational cohort study. We found that a higher genetically predicted seizure response to valproate was associated with higher valproate serum levels, indicating that the included genetic variants predispose to greater valproate exposure by affecting valproate metabolism. In addition, this higher genetically predicted response to valproate was associated with a lower risk of ischemic stroke, among valproate users only. There was no such association among individuals who were not taking valproate, and no association among lamotrigine and levetiracetam users, ruling out independent pleiotropic effects of the genetic variants or an antiepileptic drug effect. The robustness of our results is supported by replication in the MGBB biobank, an independent electronic health record database from a different continent that is representative of a clinical population. Our results support a causal effect of valproate on ischemic stroke risk and demonstrate the utility of leveraging genetic data in observational cohorts to model drug response *in silico* for drug repurposing.

Valproate’s anticonvulsant effects were discovered in 1963^27^ and it is today a commonly used drug for seizure prevention and mood stabilization.^28–30^ Valproate is a nonspecific HDAC inhibitor,^6^ and with the detection of *HDAC9* as genetic risk locus for large-artery stroke,^7, 10, 11^ the role of valproate for stroke prevention has been postulated. Various studies have investigated the association of valproate with vascular outcomes in observational studies with conflicting findings. Some have found a decreased risk of ischemic stroke^1, 2^ and myocardial infarction,^2, 3, 31, 32^ while others have found an increased risk of stroke.^33, 34^ A recent meta-analysis found an increased overall stroke risk in patients with epilepsy, but a decreased stroke risk in patients taking valproate compared to other antiepileptic drugs.^4^ Our study confirmed the presumed effect of valproate on ischemic stroke, but failed to confirm previously hypothesized associations with myocardial infarction despite better statistical power, suggesting confounding by indication or attrition bias in previous findings. To test its clinical value for ischemic stroke prevention, valproate is currently under investigation in the Sodium Valproate to Prevent Stroke (SOLVE) trial for the prevention of atherosclerosis progression in patients with large-artery stroke in the UK (ISRCTN12685153).

Our results show a decreased risk of stroke in valproate users. We were unable to investigate valproate’s effect on specific stroke subtypes because this information is not available in the UKB, limiting our insight into a specific clinical mechanism. However, we found a stronger association in individuals without a diagnosis of atrial fibrillation even though this subgroup contributed many events and thus statistical power, suggesting that valproate’s overall ischemic stroke prevention effect is not by preventing cardioembolism. The lack of association with myocardial infarction could suggest that valproate’s stroke prevention mechanism acts through other known pathways beyond slowing of atherosclerosis through *HDAC* inhibition, such as protection of the blood brain barrier,^35^ increase in ischemic tolerability through neuroprotective effects as demonstrated *in vivo* for brain ischemia, spinal cord injury and traumatic brain injury,^35–37^ inhibition of platelet aggregation,^38^ or increase in tissue plasminogen activator.^39^

In our analyses, we found the strongest effect of valproate on ischemic stroke among individuals with a history of prior stroke, despite a small cohort size. Existing data shows that post-stroke epilepsy is common, with an overall incidence of up to 7%^40^ and a high recurrence rate of up to 12% in five years^41^. If future studies confirm our observation that valproate contributes most to the prevention of recurrent ischemic stroke among stroke survivors, future trials could evaluate its utility as a treatment for post-stroke epilepsy as a dual-use secondary prevention agent.

Our study provides a compelling example of how genetic data can aid in prioritizing drug repurposing targets when observational studies are limited. The most likely reason for the conflicting evidence for the effect of valproate from observational studies is confounding by indication. As shown in our results, valproate users have a higher number of vascular risk factors compared to the general population (**Table 1**). Thus, association of valproate in solely epidemiological models use would yield an increased risk for vascular outcomes in our cohort, leading to a false conclusion from confounding bias. Our study shows that in these cases, Mendelian randomization is a powerful tool for overcoming this challenge, randomizing individuals to similar baseline characteristics **(Table 2)**, thus providing an unbiased approach that is similar to an actual clinical trial. This intriguing concept is enabled through the increasing number of studies providing genetic markers for drug response.^42^ Although valproate is a widely used and known drug, its adverse and teratogenic effects^43^ reduce its attractiveness for drug repurposing in the large collective of stroke survivors. However, our findings potentially apply to other HDAC inhibitors that have been tested *in vitro* and *in vivo*,^44^ providing further justification for stroke prevention trials employing HDAC inhibitors.

Our research illustrates the potential utility and reliability of the under-utilized primary care prescription data within the UKB. It demonstrates the capacity to generate robust and replicable findings. Our investigation yielded nearly identical estimates for the associations between the prescribed dosage of valproate and serum valproate levels within both the UKB and the MGBB, despite these data being sourced from distinct healthcare systems across two different continents, utilizing two disparate methodologies for daily intake assessment. Furthermore, we observed congruent effect estimates between the genetic score and valproate serum levels, irrespective of the absence of standardized measurements. These findings suggest that with sufficiently large cohorts, it is possible to discern true biological effects despite the diversity in electronic health care records and healthcare systems, along with their inherent weaknesses and limitations.

We acknowledge the limitations of our study. First, the SNPs used to construct the genetic score for valproate response were discovered only in individuals of European ancestry^15^ and applied to a predominantly European UKB population, raising the question whether our findings can be applied to Non-European populations. Although the leveraged genetic variants mark valproate response, they act as instrument for randomization in our study and thus it is likely that the findings also apply to individuals of non-European racial/ethnic background. Second, although we identified 2,150 valproate users, the absolute number of outcomes was low, restricting the power of our study. Third, we were not able to investigate valproate’s effect on stroke subtypes because detailed stroke phenotyping is unavailable in the UKB. Furthermore, it is possible that some of the ischemic strokes among valproate users might have been misclassified seizures with Todd’s paralysis. However, for several reasons we believe that it is unlikely that such a misclassification has substantially biased our findings: i) to drive observed associations, the majority of strokes must have been misclassified, which would raise serious concerns about the UKB phenotyping and all its associated research findings, ii) we did not find an association of the genetic response scores for lamotrigine and levetiracetam with ischemic stroke, and iii) we replicated our findings in the MGBB cohort on inpatient stroke diagnoses. Fourth, since we gathered data on valproate prescriptions, and not on valproate use, we cannot be certain that our whole cohort was in fact using valproate, but we postulate that this might only have diluted our effect estimates towards the null. Finally, our study cannot answer the question which patient collective would benefit most from valproate use, however with our limited power we found the largest effect for prevention of recurrent ischemic stroke.

In conclusion, by using an innovative Mendelian randomization approach leveraging genetic data, our study supports a causal role for valproate in the prevention of ischemic stroke, with the largest decrease in risk for recurrent ischemic stroke. Our results provide actionable evidence for the performance of clinical trials with valproate or other *HDAC* inhibitors that might have a more favorable dosing and side effect profile for the prevention of ischemic stroke.

## Disclosures

CDA has received sponsored research support from Bayer AG and has consulted for ApoPharma unrelated to this work. JR reports compensation from National Football League and Takeda Development Center Americas for consultant services, all unrelated to this work.

## Sources of Funding

CDA is supported by NIH R01NS103924, U01NS069673, American Heart Association 18SFRN34250007 and 21SFRN812095, and the Massachusetts General Hospital McCance Center for Brain Health for this work. MKG is supported by the FöFoLe program of LMU Munich (Reg.-Nr. 1120), the DFG Germany’s Excellence Strategy within the framework of the Munich Cluster for Systems Neurology (EXC 2145 SyNergy - ID 390857198), and the Fritz-Thyssen Foundation (Ref. 10.22.2.024MN) JR receives research grants from NIH and the American Heart Association-Bugher Foundation. AH received funding from the the Berta-Ottenstein programm for Advanced Clinician Scientists, Medical Faculty, University of Freiburg, Germany. SW receives funding from the German Research Foundation (WO 2385/2-1).

## Supporting information

STROBE-MR checklist

## Data Availability

UK Biobank participant data are available through the UK Biobank after approval of a research proposal. All data produced in the present work are contained in the manuscript and Supplemental Tables.

## Acknowledgments

This research has been conducted using the UK Biobank Resource under Application Number 36993. Figure 1A was created with Biorender.com.

## Notes

### Author Declarations

UK Biobank participant data are available through the UK Biobank after approval of a research proposal. The SNPs used for construction of the genetic scores were obtained from the authors of the original publication.

### Summary of Updates

To assess the robustness of our findings, we replicated observed associations in an independent cohort of valproate users of the Mass General Brigham Biobank (MGBB).

